# Clinical Laboratory Validation and Implementation of Quantitative, Real-Time PCR-based Monitoring of NPM1 Type A Mutation

**DOI:** 10.1101/19010124

**Authors:** Joelle Racchumi, Wayne Tam, Michael J Kluk

## Abstract

NPM1 is frequently mutated in acute myeloid leukemia (AML) and the mutations consist of a variety of small insertions; the most common NPM1 mutation (ie, Type A) accounts for approximately 80% of all NPM1 mutations. NPM1 mutations have been reported to have prognostic significance in AML and, more recently, monitoring mutant NPM1 levels during and after therapy has been described to predict relapse and survival. Despite the published relevance of this molecular biomarker, routine residual disease monitoring for mutant NPM1 levels has not been widely adopted in many academic clinical laboratories. In this manuscript, we present the validation and implementation of a quantitative, real-time PCR-based method for the monitoring of NPM1 Type A mutant transcripts, for use in routine practice in a clinical laboratory. We have found that this assay format is sensitive and reproducible. We describe the rigorous validation procedure and share observations that will help other clinical laboratories that may wish to implement this type of testing. We show comparison of the results from this assay to other assays and a representative case is provided which highlights the utility of the findings from this assay. Taken together, in the context of molecular testing for residual disease in AML, which is an area of practice that continues to expand, we have found that this method and protocol will serve as an appropriate option for monitoring NPM1 Type A mutation levels in clinical practice.

## INTRODUCTION

Nucleophosmin (NPM1) is a nucleolar protein expressed at high levels in many cell types which regulates several cellular functions including cell growth^1^. NPM1 is involved in tumorigenesis of different cell types through either gene rearrangement or mutation^1, 2^. Importantly, approximately 50% of acute myeloid leukemia (AML) cases with a normal karyotype (overall, approximately one third of all adult AML patients) have been shown to have mutations in NPM1^2, 3^. A variety of mutations have been reported, which consist of small insertion/deletion (in/del) frameshift variants in exon 11, frequently involving codons Trp288 or Trp290^3^. Type A mutations (c.860_863dupTCTG; p.W288Cfs*12) are the most frequent, accounting for 80% of all NPM1 mutations. These mutations alter the NPM1 protein C-terminus, and functionally, they lead to aberrant cytoplasmic localization of NPM1 in leukemic cells^3^.

NPM1 has been shown to have prognostic significance in AML that varies with co-existence of mutations in FLT3 (internal tandem duplications, ITD) and DNMT3A. In general, mutation of NPM1 in AML is associated with a favorable impact on prognosis^2^. Patients with NPM1 mutations without FLT3 ITD or DNMT3A mutations have a better prognosis than those with co-existing FLT3 ITD or DNMT3A mutations^2, 4-7^. These prognostic implications can influence the management of patients, including decisions relating to bone marrow transplantation^2, 8^. Importantly, NPM1 has been reported to be a stable molecular marker that is detectable at the time of relapse in >90% of patients with NPM1 mutant AML^8-10^.

Technically, the assessment of NPM1 mutations has been reported using both RNA-based and DNA-based methods^10, 11^. RNA-based methods have been reported to be more sensitive than DNA-based approaches and, therefore, have been used in key published monitoring studies^8, 11^. The most widely used RNA-based approach is a quantitative, real-time, reverse transcription PCR (RT-PCR) method using mutant-NPM1 specific primers where NPM1-mutant transcripts are normalized by ABL1 expression (% normalized copy number : % mutant NPM1 copies/ ABL copies).

Despite the biologic and prognostic significance of NPM1 mutations in AML, routine testing for the quantitative monitoring of NPM1 has not been widely implemented in clinical laboratories. Recently, there has been renewed interest and increasing clinical demand for monitoring mutant NPM1 levels. Ivey et al. have recently demonstrated that AML patients with persistence of mutated NPM1 transcripts in the blood after the second cycle of chemotherapy had a higher risk of relapse and a lower rate of survival than patients who did not have detectable mutated NPM1 transcripts^8^. In addition, another recent study has suggested that NPM1-mutant allele burden at diagnosis may impact prognosis in *de novo* AML^12^. The ability to detect and monitor mutant NPM1 is part of a renewed broad-based interest to employ molecular and immunophenotypic markers to detect residual /recurrent disease in acute myeloid leukemia^13, 14^.

Given these increasing clinical demands for monitoring mutant NPM1 levels, we report herein how our laboratory has validated a quantitative, real-time, RT-PCR-based method for clinical monitoring of NPM1 Type A mutation in patients with acute myeloid leukemia. Of note, this assay is distinct from the PCR/capillary electrophoresis or Next Generation Sequencing (NGS)-based methods used to screen samples for NPM1 mutations. Since the validation of this quantitative, real time, RT-PCR-based method was compliant with rigorous New York State Clinical Laboratory Evaluation Program (NY CLEP) standards, we believe this will be helpful template for other clinical laboratories that may decide to implement similar clinical, quantitative testing for NPM1, as the clinical demand for testing of this molecular biomarker continues to expand.

## MATERIALS AND METHODS

A quantitative, real-time, reverse transcription PCR (RT-PCR) assay for the detection of NPM1 Type A mutant transcripts in patient samples using RNA input from peripheral blood and/or bone marrow aspirate specimens was validated for use in our clinical laboratory. A standard one-way work flow is used between separate work areas (eg, pre-PCR area for specimen preparation, pre-PCR hood for master mix preparation, amplification area for thermocycling, and post-PCR area for detection).

Patient samples (collected in EDTA tubes) may be collected and transported briefly at room temperature, but, upon receipt, should be stored at 2C to 6C in order to prevent RNA degradation. Samples should contain at least several ml of fresh peripheral blood or bone marrow aspirates (or fresh cell preparations with >5 million cells). Red blood cell lysis and RNA extraction is performed according to standard protocol (QIAamp RNA Blood Mini Kit (Qiagen 52304)). The samples should provide at least 1 ug (1000 ng) of total RNA upon RNA extraction with A_260_/A_280_ ratios of 1.7 to 2.0.

The RNA is reverse-transcribed into cDNA according to standard protocol with 1ug (1000 ng) of total RNA input per sample in 20uL final volume (SuperScript III RT (Life Technologies, #18080085); RNaseOUT (Life Technologies, #10777019); Random Primers (Life Technologies, #48190011); dNTP mix, 10mM (Promega, #U151B); First Strand Buffer, 5x and DTT, 0.1M). Samples are incubated at 65C for 5 minutes, then placed on ice or left at 4C for at least 1 minute. Cycling condition are: 25C (×10 min), 50C (x50 min), 85C (x5 min), 4C (Hold), using an Applied Biosystems GeneAmp 9700 thermocycler or ProFlex PCR systems. cDNA samples may be kept at 4C before immediate use or stored at -20C for up to 1 week. The cDNA from the patient samples and positive and negative controls is diluted by adding 30uL of nuclease-free water (50uL total volume). 5uL of the diluted cDNA (corresponding to one tenth (100 ng RNA equivalent)) is then used as input into the quantitative, real-time PCR.

Quantitative, real-time PCR is conducted using the NPM1 mutA MutaQuant Assay (Ipsogen, Qiagen, #677513) that includes primers and probes for NPM1 Type A mutation and ABL, as well as NPM1 Type A and ABL cDNA samples for standard curve generation. Quantitative PCR is performed in an ABI 7500 Fast Real-Time PCR system (Applied Biosystems) with TaqMan Universal PCR Master Mix (Life Technologies, #4304437) using the following cycling conditions: 50C (2 min); 95C (10 min); [95C (15 sec), 60C (60 sec)] × 50 cycles. For on-board amplification plot analysis, a threshold of approximately 0.1 is set for both NPM1 and ABL. The results are reported as % Normalized Copy Number (%NCN) calculated as follows: (NPM1 Mutant A Copy Number /ABL Copy Number) × 100. The MV-4-11 cell line (negative for NPM1 Type A mutation; ATCC, CRL-9591) and the OCI-AML3 cell line (positive for NPM1 Type A mutation; DSMZ, ACC 582) may be used as control samples. An NPM1 Type A mutation low positive (LP) control near the limit of detection of the assay as well as a negative control (eg, MV-4-11 cell line) and a No Template Control (NTC) are included in every run. All samples and controls are run in duplicate reactions.

Patients tested by this assay should be previously known to have tested positive for the NPM1 Type A mutation (NPM1, NM_002520, Exon 11, c.860_863dupTCTG, p.W288Cfs*) (HG19/GRCH37) through alternate genotyping methods (e.g., NGS sequencing screening tests). Samples from patients with NPM1 mutations that are not Type A mutations are not appropriate for testing by this assay and require a separate mutation specific assay.

Myeloid NGS Panel: Targeted enrichment of 45 genes recurrently mutated in myeloid malignancies was performed using the Thunderstorm system (RainDance Technologies, Billerica, MA) using a custom primer panel followed by sequencing using the Illumina MiSeq (v3 chemistry) yielding 260-bp paired-end reads. The Myeloid NGS Panel interrogates entire coding exons or select exons, as well as flanking intron sequence for single nucleotide variants (SNV) and insertion/deletions (INDEL). Each sample is run in duplicate. The detection sensitivity of the assay for SNV and INDEL, as determined by a comprehensive validation, is approximately 2% for SNV, and 1% for INDEL.

This work is covered under the Weill Cornell IRB Protocol #: 1007011151.

## RESULTS

This quantitative, real-time, RT-PCR assay underwent full validation compliant with NY State CLEP standards. The accuracy of the assay was assessed by comparing real time RT-PCR results to orthogonal methods, including next generation sequencing and digital droplet PCR (ddPCR); the accuracy was 100% for both 10 positive and 10 negative samples tested by alternate methods (see Table 1 and Supplemental Table 1). The Inter-run and Intra-run reproducibility of the assay showed minimal variation (ie, ≤0.5 log_10_) for several different negative samples, and several positive samples, including both high level and low level positive samples, near the lower limit of detection (see Table 2 and Supplemental Table 2). In addition, the NPM1 Type A Mutation and ABL1 standard curves also showed excellent reproducibility between runs with average slopes of -3.45, -3.47 and R^2^ values of 1.00 (Supplemental Figures 1 and 2). In terms of sensitivity, the maximal reproducible analytic sensitivity of this assay was approximately 0.01% NCN (NPM1 MutA /ABL) (0.006%-0.013%) demonstrated by patient sample and cell line serial dilutions (see Table 3 and Supplemental Table 3). In addition, this RT-PCR assay showed excellent concordance of the %NCN values with a separate ddPCR assay across the range of values in a dilution series and for several samples tested near the lower limit of detection (see Table 4).

**Table 1:** Accuracy for Positive Samples. Real Time PCR Compared to Reference Method(s)

| Sample # | Specimen | NPM1 Type A Mutant RT-PCR Result | NPM1 Type A Mutant RT-PCR (% , NCN) | Next Gen. Seq. Reference Result | ddPCR Reference Result (if available) (% , NCN) | Path. Diagnosis |
| --- | --- | --- | --- | --- | --- | --- |
| P1 | PB | Positive | 216.28 | Positive, c.860_863dupTCTG; p.W288Cfs*12, Type A, 56% VAF | Positive | AML |
| P2 | PB | Positive | 250.59 | Positive, c.860_863dupTCTG; p.W288Cfs*12, Type A, 40% VAF | Positive | AML |
| P3 | BM | Positive | 179.06 | Positive, c.860_863dupTCTG; p.W288Cfs*12, Type A, 28% VAF | Positive | AML |
| P9 | PB | Positive | 263.29 | Positive, c.860_863dupTCTG; p.W288Cfs*12, Type A, 51% VAF | Positive | AML |
| P11 | PB | Positive | 620.63 | Positive, c.860_863dupTCTG; p.W288Cfs*12, Type A, 47 % VAF | Positive | AML |
| P13 | PB | Positive | 405.03 | Positive, c.860_863dupTCTG; p.W288Cfs*12, Type A, 50% VAF | N/A | AML |
| P14 | PB | Positive | 82.44 | Positive, c.860_863dupTCTG; p.W288Cfs*12, Type A, 43% VAF | Positive | AML |
| P16 | PB | Positive | 577.4 | Prior AML sample with NPM1 Mutant Type A (c.860_863dupTCTG, 46% VAF). Concurrent sample: AML (87% Blasts). | Positive | AML |
| P18 | PB | Positive | 83.94 | History of NPM1 Mutant Type A (c.860_863dupTCTG; 48% VAF). Flow Cytometry (within 9 d of current sample) showed Residual AML: 1% Blasts. | Positive | Residual AML |
| P19 | PB | Positive | 398.09 | History of NPM1 Mutant Type A (c.860_863dupTCTG) NGS and Pathology (within 12 d of current sample) showed AML with NPM1 Type A Mutation (18% VAF). | Positive | Residual AML |

NOTES: %NCN = %Normalized Copy Number = (NPM1 Mutant Type A Copies/ ABL Copies)\*100

**Table 2:**
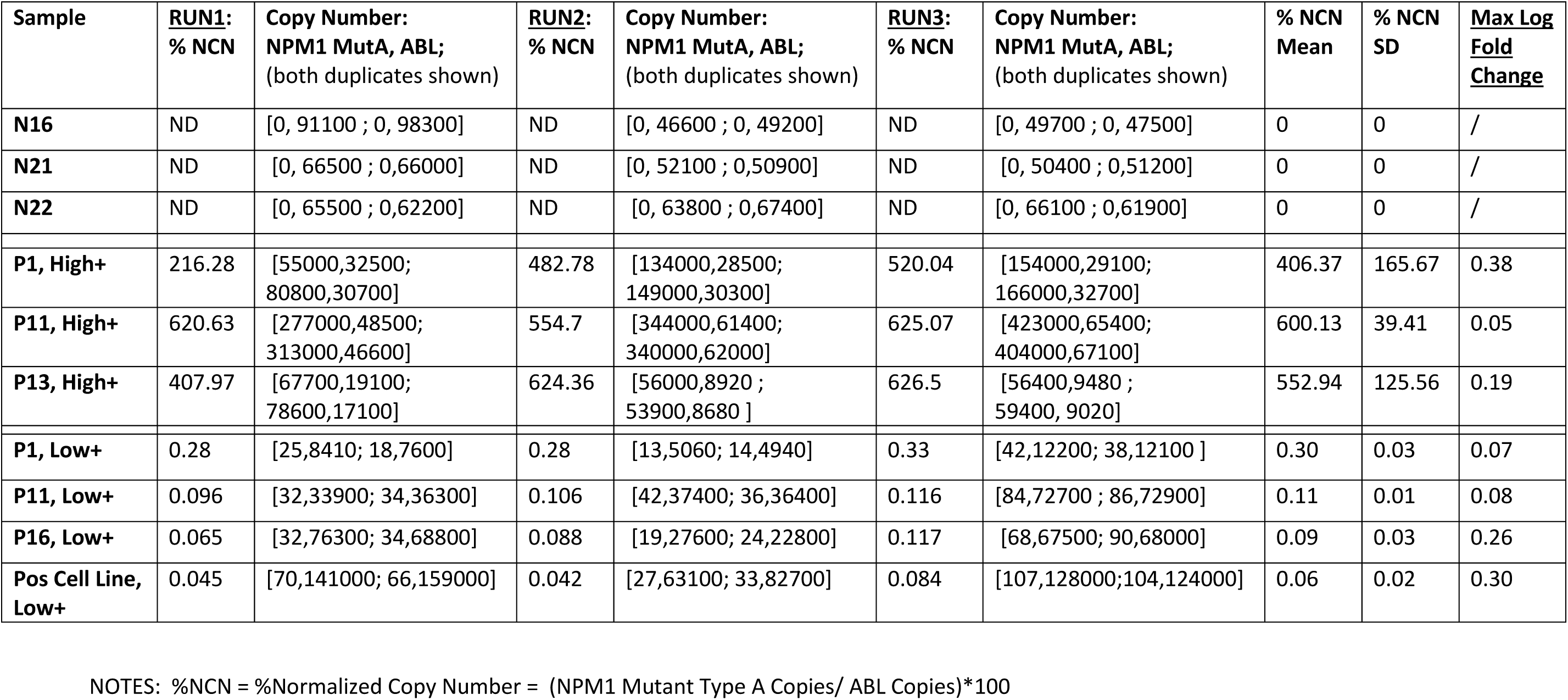
Inter-Run Reproducibility.

| **Sample** | **<u>RUN1:</u>**<br>**% NCN** | **Copy Number:**<br>**NPM1 MutA, ABL;**<br>**(both duplicates shown)** | **<u>RUN2:</u>**<br>**% NCN** | **Copy Number:**<br>**NPM1 MutA, ABL;**<br>**(both duplicates shown)** | **<u>RUN3:</u>**<br>**% NCN** | **Copy Number:**<br>**NPM1 MutA, ABL;**<br>**(both duplicates shown)** | **% NCN**<br>**Mean** | **% NCN**<br>**SD** | **<u>Max Log</u>**<br>**<u>Fold</u>**<br>**<u>Change</u>** |
| --- | --- | --- | --- | --- | --- | --- | --- | --- | --- |
| **N16** | ND | [0, 91100 ; 0, 98300] | ND | [0, 46600 ; 0, 49200] | ND | [0, 49700 ; 0, 47500] | 0 | 0 | / |
| **N21** | ND | [0, 66500 ; 0,66000] | ND | [0, 52100 ; 0,50900] | ND | [0, 50400 ; 0,51200] | 0 | 0 | / |
| **N22** | ND | [0, 65500 ; 0,62200] | ND | [0, 63800 ; 0,67400] | ND | [0, 66100 ; 0,61900] | 0 | 0 | / |
| **P1, High+** | 216.28 | [55000,32500;<br>80800,30700] | 482.78 | [134000,28500;<br>149000,30300] | 520.04 | [154000,29100;<br>166000,32700] | 406.37 | 165.67 | 0.38 |
| **P11, High+** | 620.63 | [277000,48500;<br>313000,46600] | 554.7 | [344000,61400;<br>340000,62000] | 625.07 | [423000,65400;<br>404000,67100] | 600.13 | 39.41 | 0.05 |
| **P13, High+** | 407.97 | [67700,19100;<br>78600,17100] | 624.36 | [56000,8920 ;<br>53900,8680 ] | 626.5 | [56400,9480 ;<br>59400, 9020] | 552.94 | 125.56 | 0.19 |
| **P1, Low+** | 0.28 | [25,8410; 18,7600] | 0.28 | [13,5060; 14,4940] | 0.33 | [42,12200; 38,12100 ] | 0.30 | 0.03 | 0.07 |
| **P11, Low+** | 0.096 | [32,33900; 34,36300] | 0.106 | [42,37400; 36,36400] | 0.116 | [84,72700 ; 86,72900] | 0.11 | 0.01 | 0.08 |
| **P16, Low+** | 0.065 | [32,76300; 34,68800] | 0.088 | [19,27600; 24,22800] | 0.117 | [68,67500; 90,68000] | 0.09 | 0.03 | 0.26 |
| **Pos Cell Line,**<br>**Low+** | 0.045 | [70,141000; 66,159000] | 0.042 | [27,63100; 33,82700] | 0.084 | [107,128000;104,124000] | 0.06 | 0.02 | 0.30 |

NOTES: %NCN = %Normalized Copy Number = (NPM1 Mutant Type A Copies/ ABL Copies)\*100

**Table 3:** Analytic Sensitivity. Patient Sample.

| Table 3: Analytic Sensitivity. Patient Sample. |  |  |  |  |  |
| --- | --- | --- | --- | --- | --- |
| Patient Sample Dilution<br>(% Pos Control RNA) | NPM Type A Mutant,<br>Number of Copies | ABL, Number of Copies | % NCN,<br>NPM1 Type A Mutant/ABL | Mean,<br>% NCN | SD |
| P11_100% | 121808.750 | 35411.234 | 343.983 | 355.525 | 16.323 |
| P11_100% | 131026.953 | 35695.621 | 367.067 |  |  |
| P11_10% | 27605.953 | 50782.969 | 54.361 | 51.901 | 3.479 |
| P11_10% | 27324.826 | 55267.434 | 49.441 |  |  |
| P11_1% | 2317.520 | 43984.738 | 5.269 | 5.610 | 0.483 |
| P11_1% | 2429.558 | 40819.695 | 5.952 |  |  |
| P11_10-1% | 342.882 | 40465.391 | 0.847 | 0.822 | 0.036 |
| P11_10-1% | 322.556 | 40501.297 | 0.796 |  |  |
| P11_10-2% | 58.689 | 56396.590 | 0.104 | 0.100 | 0.006 |
| P11_10-2% | 54.337 | 57233.090 | 0.095 |  |  |
| P11_10-3% | **13.156** | **94619.969** | **0.014** | **0.013** | **0.002** |
| P11_10-3% | **11.611** | **98536.828** | **0.012** |  |  |
| P11_10-4% | 1.466 | 88979.555 | 0.002 | 0.001 | 0.001 |
| P11_10-4% | 0.110 | 90285.188 | 0.000 |  |  |
| P11_10-5% | 0.000 | 96103.969 | 0.000 | 0.000 | 0.000 |
| P11_10-5% | 0.004 | 87504.039 | 0.000 |  |  |
| N14 | 0.000 | 84404.031 | 0.000 | 0.000 | 0.000 |
| N14 | 0.000 | 75076.672 | 0.000 |  |  |

**Table 4:** Real Time PCR vs. ddPCR.

| **Table 4: Real Time PCR vs. ddPCR** |  |  |
| --- | --- | --- |
| **Sample** | **Real Time PCR,**<br>**%NCN, (NPM1 Type A Mutant/ABL)** | **dd PCR,**<br>**%NCN, (NPM1 Type A Mutant/ABL)** |
| P11_100% | 355.5253 | 255.9524 |
| P11_10% RNA Dilution | 51.9009 | 36.5452 |
| P11_1% RNA Dilution | 5.6104 | 2.7967 |
| P11_10-1% RNA Dilution | 0.8219 | 0.4374 |
| P11_10-2% RNA Dilution | 0.0995 | 0.0593 |
| P11_10-3% RNA Dilution | 0.0128 | 0.0085 |
| **Sample** | **Real Time PCR,**<br>**%NCN, (NPM1 Type A Mutant/ABL)** | **dd PCR,**<br>**%NCN, (NPM1 Type A Mutant/ABL)** |
| P1_RNA Dilution_Run1 | 0.2765 | 0.3099 |
| P1_RNA Dilution_Run2 | 0.2769 | 0.4058 |
| P11_RNA Dilution_Run1 | 0.0961 | 0.0719 |
| P11_RNA Dilution_Run2 | 0.106 | 0.0807 |
| P16_RNA Dilution_Run1 | 0.0651 | 0.1411 |
| P16_RNA Dilution_Run2 | 0.0879 | 0.1216 |
| P_Sample A_Run1 | 0.043 | 0.027 |
| P_Sample A_Run2 | 0.052 | 0.025 |

The results from a representative patient aid in the demonstration of how this testing can be helpful in the setting of routine clinical practice. The patient is a 39 year old woman who was recently diagnosed with acute myeloid leukemia; cytogenetic analysis of which revealed a normal female karyotype. Molecular testing by NGS at the time of diagnosis was positive for a FLT3 internal tandem duplication (39 bp insert) and was also positive for the NPM1 Type A mutation (c.860_863dupTCTG; p.W288Cfs*12, 42% VAF), as well as variants in DNMT3A (c.2171A>G; p.Y724C, 51% and c.932_945delTGTCTTGGTGGATG; p.V311Dfs*8, 51%) and NF1 (c.7789T>C; p.S2597P, 52%). The patient was treated with 7+3 induction chemotherapy, including midostaurin, and had a bone marrow biopsy on day 24. The bone marrow biopsy (Figure 1) revealed a hypercellular marrow with myeloid hyperplasia and left shifted myeloid maturation with 6% blasts as well as megakaryocytic hyperplasia. No circulating blasts were observed in the peripheral blood. Given that the patient had recently received growth factors, it was not entirely clear if the morphologic findings were a result of recent growth factor administration, or whether they represented impending recurrence of the acute myeloid leukemia. Likewise, flow cytometry was not definitive for residual disease. Since the patient was known to have an NPM1 Type A mutation, a peripheral blood sample was tested by the NPM1 Type A mutation RT PCR assay described above, and the patient was found to have a high level of NPM1 Type A mutation (63.3% NCN, NPM1 Type A Mutation/ABL; Figure 1). Next generation sequencing with the Myeloid NGS Panel was performed on the genomic DNA from the peripheral blood sample and also revealed the Type A NPM1 mutation (0.4% variant allele frequency (VAF); Figure 1). The prior FLT3 ITD (39bp) was also noted on NGS (0.4% VAF) and the prior DNMT3A variants were also seen at low VAF (DNMT3A c.2171A>G; p.Y724C, 1.5% VAF and c.932_945delTGTCTTGGTGGATG; p.V311Dfs*8, 3.1%). 14 days after the NPM1 Type A mutation RT-PCR result, a follow up peripheral blood specimen was subsequently sent for flow cytometry and revealed that the patient had 25% circulating abnormal blasts, consistent with residual/recurrent acute myeloid leukemia. This aggressive clinical expansion of the abnormal blasts was consistent with the high level of mutant NPM1 mutation detected by the RT-PCR assay.

**Figure 1:**
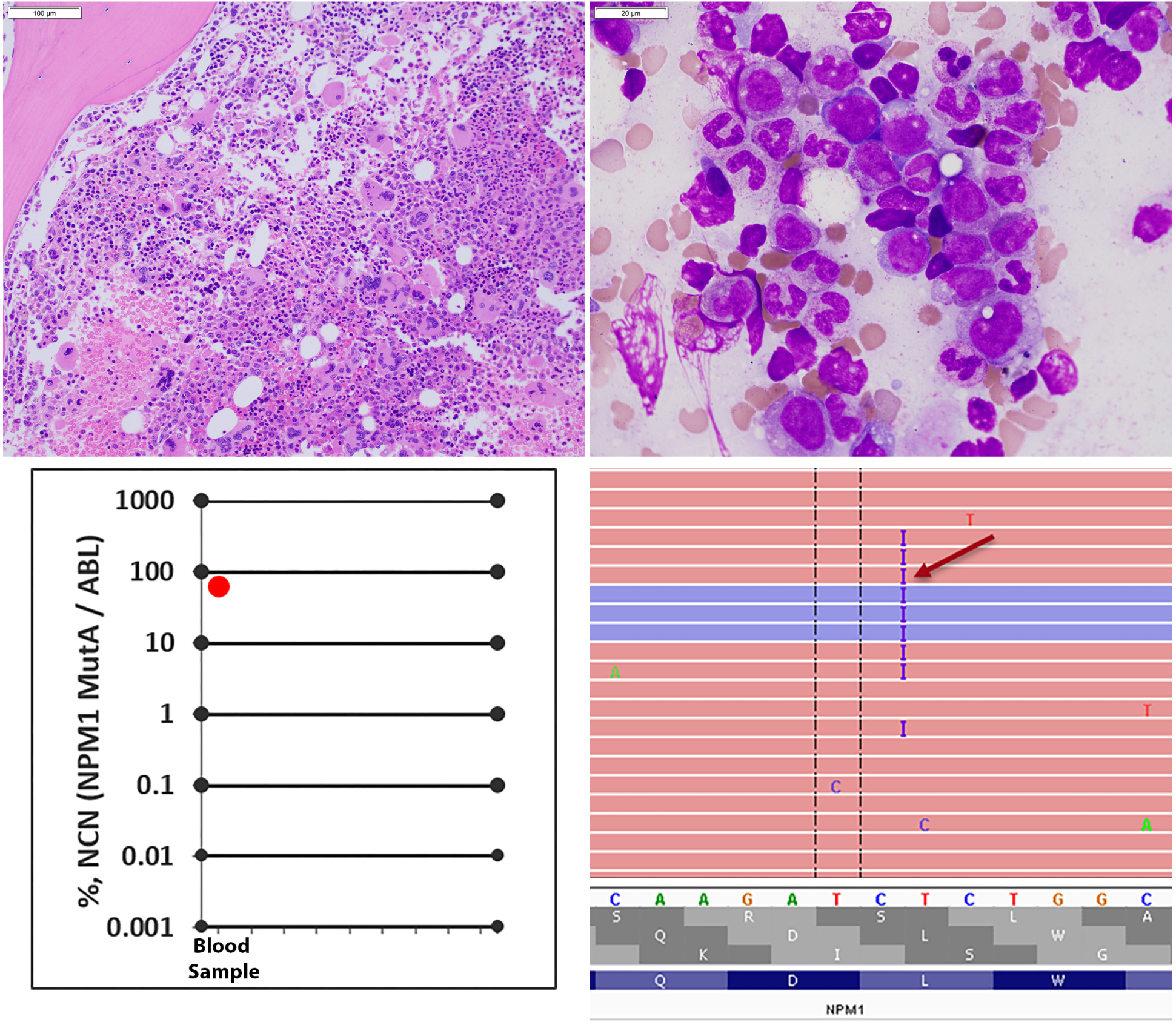
Data from a Representative Patient. An image of the H&E stained core biopsy (upper left) shows overall hypercellularity and relative myeloid hyperplasia. An image of the Wright-Giemsa stained bone marrow aspirate smear (upper right) shows maturing myeloid elements with full maturation, but with a slight left in maturation. Graph of the NPM1 Type A mutation Real Time, RT-PCR results (lower left) showing the presence of the NPM1 Type A mutation (63.3% NCN, Normalized Copy Number: NPM1 Type A Mutation/ABL). Image of the sequencing reads (lower right) in Integrative Genomics Viewer (IGV©, Broad Institute) from genomic DNA-based sequencing also reveals the NPM1 Type A mutation (0.4% variant allele frequency).

Given the difference in the NPM1 Type A mutation levels observed between this real time, RT-PCR method (using RNA input) and the next generation sequencing (NGS) method (using genomic DNA input) for the representative case described above, we compared the NPM1 Type A mutation levels as measured by the RT-PCR method to mutation levels determined by NGS (using genomic DNA-based input) when material from the same sample was available for both assays. The results are summarized in Table 5 and demonstrate the superior sensitivity of the RNA-based RT-PCR method for NPM1 Type A mutation detection compared to genomic DNA-based Myeloid NGS panel method, even given abundant, robust read depths (eg, 1500-2000) at this NPM1 location by the NGS assay. This greater sensitivity of the RNA-based RT-PCR method is consistent with prior reports indicating the greater sensitivity of RNA-based methods compared to genomic DNA-based input for the detection of mutant NPM1^11, 14^. This finding is also consistent with the fact that NPM1 is known to be a highly expressed transcript/protein^15-17^.

**Table 5:** Real Time PCR (RNA input) vs. 45 Gene-Myeloid NGS Panel (Genomic DNA input)

|  |  |  |  |
| --- | --- | --- | --- |
| Case | Real Time, RT-PCR<br>NPM1 MutA/ABL, %NCN | Myeloid NGS Panel,<br>%VAF | Myeloid NGS Panel,<br>(Mutant Reads/Total Reads) |
| 1 | 0.025 | 0% | 0/1357, 0/1972 |
| 2 | 0.049 | 0% | 0/2462, 0/2001 |
| 3 | 57.21 | 0.9% | 20/1694, 12/1810 |
| 4 | 0 | 0% | 0/1297, 0/1189 |
| 5 | 0.015 | 0% | 0/2020, 0/1935 |
| 6 | 65.98 | 0.8% | 2/102, 0/145 |
| 7 | 63.26 | 0.4% | 12/2451, 6/2226 |
| 8 | 0.0065 | 0% | 0/1572, 0/1308 |

Taken together, our validation studies demonstrate that this quantitative, real-time, RT-PCR method using RNA (ie, cDNA) input is a sensitive and reproducible method for the detection of Type A mutant NPM1 transcripts in patient samples. In addition, the representative case shows how this assay can provide important findings when used in the routine clinico-pathological workup of patients being monitored for residual acute myeloid leukemia. Lastly, consistent with prior understanding, we have found that this assay format provides greater sensitivity than genomic DNA based approaches (eg, 45 gene Myeloid NGS panel).

## DISCUSSION

Residual disease monitoring is becoming increasingly adopted in routine clinical care for acute myeloid leukemia (AML); flow cytometric methods focus on detecting abnormal myeloid cells with altered immunophenotypic profiles, but this approach has some limitations due to the heterogeneity of immunophenotypic abnormalities in AML, as well as technical challenges^14^. Molecular methods, which can complement flow cytometric-based studies, are increasingly being implemented as an approach for monitoring residual disease in AML. Although AML cases frequently harbor at least 1 or 2 driver mutations^18^, not all of these mutations are suitable for residual disease monitoring^14, 19, 20^.

NPM1 mutations in AML were described approximately 15 years ago^3^ and the mechanism of pathogenesis continues to be studied^21^. Meanwhile, the utility of tracking mutant NPM1 as a reliable marker of disease status in AML continues to expand^8, 10, 13, 14^. Herein, we report the validation and clinical implementation of a quantitative, real-time, RT-PCR assay for the detection and monitoring of the most common NPM1 mutation (ie, Type A NPM1 mutation) in the routine clinical management of patients with AML or other myeloid disorders. We have found that this assay is a sensitive (ie, maximal reproducible sensitivity 0.01% NCN), reproducible (ie, 100% concordance with known negative and known positive samples, ≤0.5 log_10_-fold variation between replicates/batches) platform for the detection and monitoring of the NPM1 Type A mutation.

Based on our experience, the following observations may be helpful to other laboratories who wish to implement this assay. In terms of sample requirements, we have found that it is important to receive 1-2 EDTA tubes each containing several mls of peripheral blood or bone marrow aspirate, and that RNA extraction and elution should be optimized to permit appropriate RNA concentrations, so that the requisite RNA input (1000 ng) can be input into the cDNA synthesis reaction. This is important since it will influence the overall sensitivity of the assay, by impacting the number of ABL copies obtained (ideally, at least 10,000 ABL copies obtained per replicate reaction, *see below for additional details*). In terms of results interpretation, a NTC (No Template Control) sample, which is included in each batch of tests, should be negative for both NPM1 Type A and ABL. A negative control sample (eg, MV411 cell line or other known negative sample) should be negative for NPM1 Type A mutation, but positive for ABL. A low positive control sample should also be included in each batch of testing and should be positive for NPM Type A mutation, (eg, near the lower limit of the reportable range of the assay, 0.1% NCN), and should also be positive for ABL. The standard curves (NPM1 Mut A and ABL) should each show slopes of -3.3 to -3.6 and R2 values of ≥ 0.95. The data obtained should be consistent between duplicates such that the CT (cycle threshold) value in between duplicates is +/-1 to 2 cycles. For patient samples, NPM1 Type A is considered negative if it is not detected in both duplicate reactions and the replicates show at least 10,000 ABL copies. NPM1 Type A is considered positive if it is detectable in both duplicate reactions with at least 10 NPM1 mutA copies in at least 10,000 ABL Copies (ie, ≥ 0.1% NCN (NPM1 mutA/ABL). If one of the duplicates has a positive value for NPM1 mutA and the other replicate is not detected, or if both duplicates show a value of <0.1% NCN (NPM1 mutA/ABL) or <10 NPM1 mutA copies, then new cDNA is prepared and the assay is repeated. If the repeat reactions yield a positive result in both of the new replicates, then the result will be considered positive. Similarly, if the repeat reactions yield a negative result in both of the new replicates, then the result will be considered negative. Lastly, if one of the new replicates is low positive and then the other new replicate is negative, then the average of the positive replicates from the first and second runs are taken, and the result is considered positive. For patients with results <0.1% NCN (NPM1 mutA/ABL), a note can be included indicating that it represents a borderline result. In addition, if a patient sample is negative for NPM1 Type A, but the ABL copy number is less than 10,000, then the sample is considered suboptimal for minimal residual disease, and a note can be added to the report to suggesting testing a new sample for further evaluation. Lastly, as is true for all molecular testing, interpretation of the results in the context of the clinical and pathologic findings (eg, flow cytometry residual disease testing, etc) is helpful.

According to European Leukemia Net MRD Working Party (ELN) consensus document^14^, a sensitivity of 0.1% is suggested for residual disease platforms, and real-time qPCR platforms with cDNA-based inputs are recommended because of their sensitivity. Recommended sampling time points include at diagnosis, after 2 cycles of standard induction/consolidation chemotherapy and at the end of treatment, using peripheral blood and bone marrow samples. In addition, after the end of treatment, molecular MRD assessment is recommended every 3 months for 24 months. Upon serial sampling, molecular progression has been proposed to be defined as an increase in copy numbers of 1 log_10_ or more between any 2 positive samples.

One of the limitations of the current assay is that it is mutation-specific and, therefore, is not used to detect and monitor “Non-Type A” mutations in NPM1, which would require different assays. Therefore, for monitoring of NPM1 Type A mutation levels with this assay, the patient should be known to have had the NPM1 Type A mutation (c.860_863dupTCTG; p.W288Cfs*12) at initial diagnosis, typically identified through other testing (e.g., Myeloid Gene Next Generation Sequencing (NGS) Panel). The overall clinical sensitivity, for this assay is 75-80% given that among all patients with NPM1 mutations, the Type A NPM1 mutation accounts for 75-80% of all NPM1 mutations and this assay is used to detect only NPM1 Type A mutations. Since the NPM1 Type A mutation primers and probes may show unpredictable cross-reactivity against other types of NPM1 mutations, patients with “Non-Type A” NPM1 mutations are not tested by this assay; those patients will require testing by separate mutation specific primer sets which were not part of this assay validation. Recently, a multiplex digital droplet platform capable of detecting and quantitating several different NPM1 mutation types in one assay has been described, although in that assay platform, the type of NPM1 mutation detected in a sample is not specifically identified. As technology evolves, NGS-based and digital droplet PCR-based methods will likely be increasingly integrated into MRD testing algorithms, especially as the need for tracking several potential molecular biomarkers of disease becomes increasingly desired. For example, the Myeloid NGS assay used for comparison here, was a “typical” Myeloid NGS panel (ie, 45 gene panel), similar to those often used in academic centers and commercial laboratories for testing at diagnosis, which are often amplification-based sequencing assays, without unique molecular barcodes and consist of multigene panels (eg, 40-50 genes). As technology advances, the use of smaller NGS panels that incorporate error reduction measures (eg, unique molecular barcodes) and provide ultrahigh sequencing depths that permit very sensitive detection of select consensus “molecular MRD biomarkers” in myeloid diseases will likely expand. Indeed, such approaches have recently been published for NPM1^22^ with “ultradeep” NGS sequencing (eg, 300,000 sequencing depths) using genomic DNA input. The advantages of DNA-based NGS methods include the stability DNA as an input substrate and the ability to interrogate multiple molecular biomarkers simultaneously in a single assay. However, potential disadvantages of such NGS approaches, at this point in time, include the limited availability of informatics support for such “ultradeep” NGS assay platforms, the higher amount of input DNA required, the expense of NGS, and the overall complexity of NGS assays, which require appropriate, dedicated institutional resources.

Overall, a testing strategy for AML is developing whereby several assays are performed at diagnosis (including NGS panel testing, cytogenetic analysis as well as rapid, focused assays interrogating potential therapeutic targets (eg, FLT3)), which is then followed (ie, during and post-therapy) by sequential monitoring of mutations known to be reliable markers of residual disease (eg, NPM1). In this respect, the assay described herein offers an effective option for serial testing of patients with myeloid diseases that are positive for the NPM1 Type A mutation.

## Data Availability

N/A

